# Clinical assessment and validation of a rapid and sensitive SARS-CoV-2 test using reverse-transcription loop-mediated isothermal amplification

**DOI:** 10.1101/2020.05.12.20095638

**Authors:** Melis N. Anahtar, Graham E.G. McGrath, Brian A. Rabe, Nathan A. Tanner, Benjamin A. White, Jochen K.M. Lennerz, John A. Branda, Constance L. Cepko, Eric S. Rosenberg

**Affiliations:** Department of Pathology, Massachusetts General Hospital and Harvard Medical School, Boston, MA 02114; Department of Medicine, Massachusetts General Hospital, Boston, MA 02114; Departments of Genetics and Ophthalmology, Harvard Medical School, Boston, MA 02115; New England Biolabs, Ipswich, MA 01938; Department of Emergency Medicine, Massachusetts General Hospital, Boston, MA 02114; Howard Hughes Medical Institute, Chevy Chase, MD 20815; Blavatnik Institute, Boston, MA 02115

## Abstract

Amid the enduring COVID-19 pandemic, there is an urgent need for expanded access to rapid and sensitive SARS-CoV-2 testing worldwide. Here we present a simple clinical workflow that uses a sensitive and highly specific colorimetric reverse-transcription loop-mediated isothermal amplification (RT-LAMP) to detect SARS-CoV-2 and takes forty minutes from sample collection to result. This test requires no specialized equipment and costs a few dollars per sample. Nasopharyngeal samples collected in saline were added either directly (unprocessed) to RT-LAMP reactions or first inactivated by a combined chemical and heat treatment step to inhibit RNases and lyse virions and human cells. The specimens were then amplified with two SARS-CoV-2-specific primer sets and an internal specimen control; the resulting color change was visually interpreted. While direct addition of unprocessed specimens to RT-LAMP reactions could reliably detect samples with abundant SARS-CoV-2, the assay sensitivity markedly increased after the addition of an inactivation step. In 62 clinical samples with a wide range of SARS-CoV-2 nucleic acid concentrations, the assay had 87.5% sensitivity and 100% specificity with a limit of detection at least 25 copies/μL, making it an ideal test to rule in infection. To increase sensitivity, samples that tested negative for SARS-CoV-2 by direct sample addition could be reflexed to a purification step, to increase the effective per-reaction sample input volume. In 40 purified samples, the assay yielded a 90% sensitivity and 100% specificity, with a limit of detection comparable to commercially available real-time PCR-based diagnostics that have received Emergency Use Authorization (EUA) from the FDA. This test for SARS-CoV-2 can be performed in a range of settings for a fraction of the price of other available tests, with limited equipment, and without relying on over-burdened supply chains to increase overall testing capacity.

## Introduction

The worldwide spread of COVID-19 has led to an unprecedented need for rapid, accurate, affordable, and readily available SARS-CoV-2 tests. Over fifty molecular assays for the detection of SARS-CoV-2 RNA have received FDA EUA to date, but they have not met the need for widespread testing demand due to several critical factors including: a high cost per reportable result (in the 15-40 US dollar range), and costly up-front capital equipment such as proprietary testing platforms, real-time amplification and detection platforms, or automated RNA extraction equipment and consumables that are in limited supply. In general, these tests must be performed by highly trained molecular laboratory professionals, in well-resourced laboratories. The development of more simple, rapid, and low-cost diagnostics that do not rely on the same supply chains as other COVID-19 tests could help rapidly and substantially expand testing capabilities.

Alternative rapid tests under development to detect SARS-CoV-2 rely on detection of viral antigen using lateral-flow immunoassays (LFA). While extremely convenient, respiratory viral LFAs tend to be less sensitive than nucleic-acid amplification methods, with an average sensitivity of 61-75% [1], and require high affinity, non-cross-reacting antibodies that are arduous to isolate. As an alternative to less sensitive antigen detection methods and difficult or expensive real-time PCR tests, isothermal amplification methods such as loop-mediated isothermal amplification (LAMP) and recombinase polymerase amplification (RPA) enable sensitive detection of nucleic acids with just the use of a stable heat source in as little at 15 minutes. Colorimetric RT-LAMP expands on the basic LAMP technology with a one-pot reaction that contains both reverse transcriptase and DNA polymerase with visual detection of nucleic acid amplification due to a pH indicator dye within the master mix, obviating the need for additional detection equipment [2]. LAMP has been used to detect many pathogens including Zika virus [3], *Mycobacterium tuberculosis* [4], malaria [5], and human leishmaniasis [6]. RT-LAMP has also been performed on extracted RNA for subsequent CRISPR-Cas12-based detection of SARS-CoV-2 [7].

To develop a truly accessible sample-to-answer nucleic acid-based diagnostic test, one must couple a simple detection method with an equally simple sample preparation method. The simplest sample preparation method is to directly add sample to the amplification reaction, but this can be problematic for several reasons. Endogenous RNases present in body fluids can degrade target RNA and infectious virus contained in the sample may increase the risk of laboratory acquired infection among technologists who handle the specimens. Specifically, with colorimetric RT-LAMP, the buffer and phenol red in universal viral transport media (UTM), one of the three collection media recommended by the Centers for Disease Control for SARS-CoV-2 testing, may interfere with the pH-mediated color change. To circumvent these issues, most SARS-CoV-2 molecular tests use extracted RNA as input, but RNA extraction is expensive, time consuming, laborious, and extraction kits are in short supply.

As part of an ongoing quality improvement initiative, we tested 135 clinical nasopharyngeal samples collected from Massachusetts General Hospital patients who were admitted or evaluated in the Emergency Department during the COVID-19 pandemic to determine the testing characteristics of three diagnostic strategies using colorimetric RT-LAMP. The first was the direct-from-sample approach, including samples collected in either universal transport media or sterile physiologic saline. The second incorporated an upfront five-minute chemical and heat inactivation step to inhibit RNases and lyse virions. The third strategy incorporated an additional nucleic acid purification step using a solution of silica particles (“glass milk”) to increase the effective sample input volume into the RT-LAMP reaction. Regardless of the sample preparation method, each sample was amplified with two SARS-CoV-2-specific primer sets for the ORF1a gene [8] and N gene [9]. An additional primer set for the human actin B gene also served as an internal specimen control to detect the presence of inhibitors. A negative and positive control were tested with every set of clinical samples. After the thirty-minute heating step, the results were visually interpreted (Fig. 1 and Table 1.)

**Figure 1.**
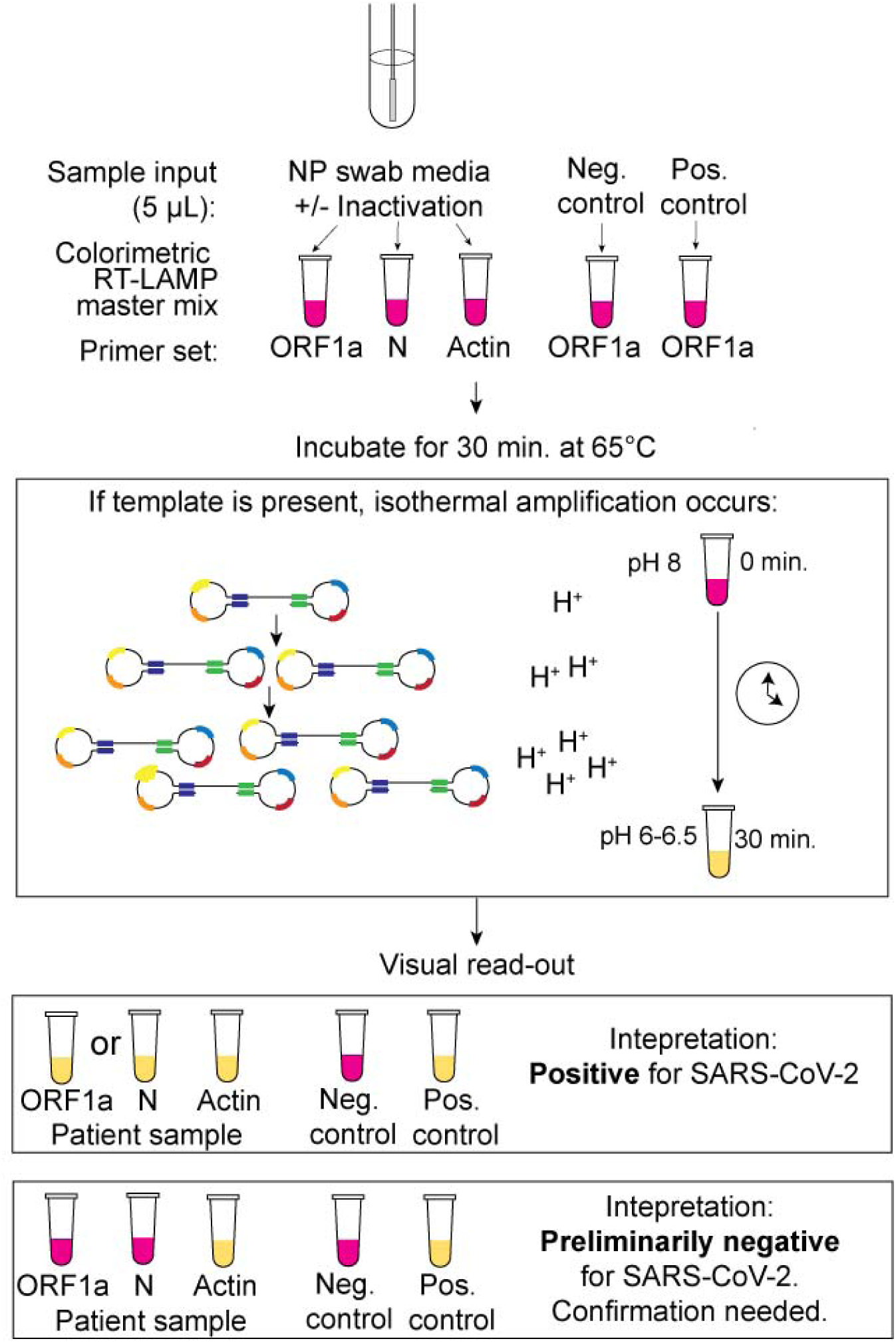
Schematic for the use of reverse-transcription loop-mediated isothermal amplification (RT-LAMP) directly from nasopharyngeal (NP) specimen using two SARS-CoV-2 specific primers, which target the ORF1a and N genes, and one internal specimen control targeting the human actin gene. Prior to sample addition to the RT-LAMP reaction, the NP specimen can undergo a five-minute heat and chemical inactivation step to destroy endogenous RNases and lyse viral particles and human cells. The RT-LAMP reaction occurs at 65 °C for 30 minutes, during which the amplification of SARS-CoV-2 RNA generates protons that decrease the pH of the reaction mix and result in a color change due to the media’s colorimetric pH indicator. Samples are removed from the heat block, immersed in ice to enhance the color brightness, and color change is visually determined. If the controls are valid, a yellow color change with the ORF1a and/or N gene primers indicates the presence of SARS-CoV-2 RNA in the sample.

**Table 1.** Interpretation matrix.

| RT-LAMP<br>Primer Set Result |  |  | Action |
| --- | --- | --- | --- |
| N | ORF1a | Actin |  |
| + | + | ± | Report “**Positive** for SARS-CoV-2 (COVID-19).” |
| - | + | ± | Report “**Positive** for SARS-CoV-2 (COVID-19).” |
| + | - | ± | Report “**Positive** for SARS-CoV-2 (COVID-19).” |
| - | - | + | Reflex to qPCR or glass milk purification. |
| - | - | - | **Invalid** result.<br>Report: “Specimen re-collection and submission is recommended.” |
| Ambiguous color change |  | + | **Indeterminant** result. Repeat test. |

## Results

### Comparison of UTM versus saline transport media

With the goal of directly adding unprocessed sample into the RT-LAMP reaction (Fig. 2A), we first optimized the transport media input volume. We added increasing amounts of transport media to a standardized reaction containing 1,000 copies of SARS-CoV-2 control RNA. UTM interfered with the colorimetric readout, with complete inhibition of the pH-mediated color change with 3 μL of input, while saline had little effect (Fig. 2B). Subsequent experiments were conservatively performed with 1 μL of UTM and 5 μL of saline sample input to facilitate robust assay performance in the setting of intrinsic clinical sample variability.

**Figure 2.**
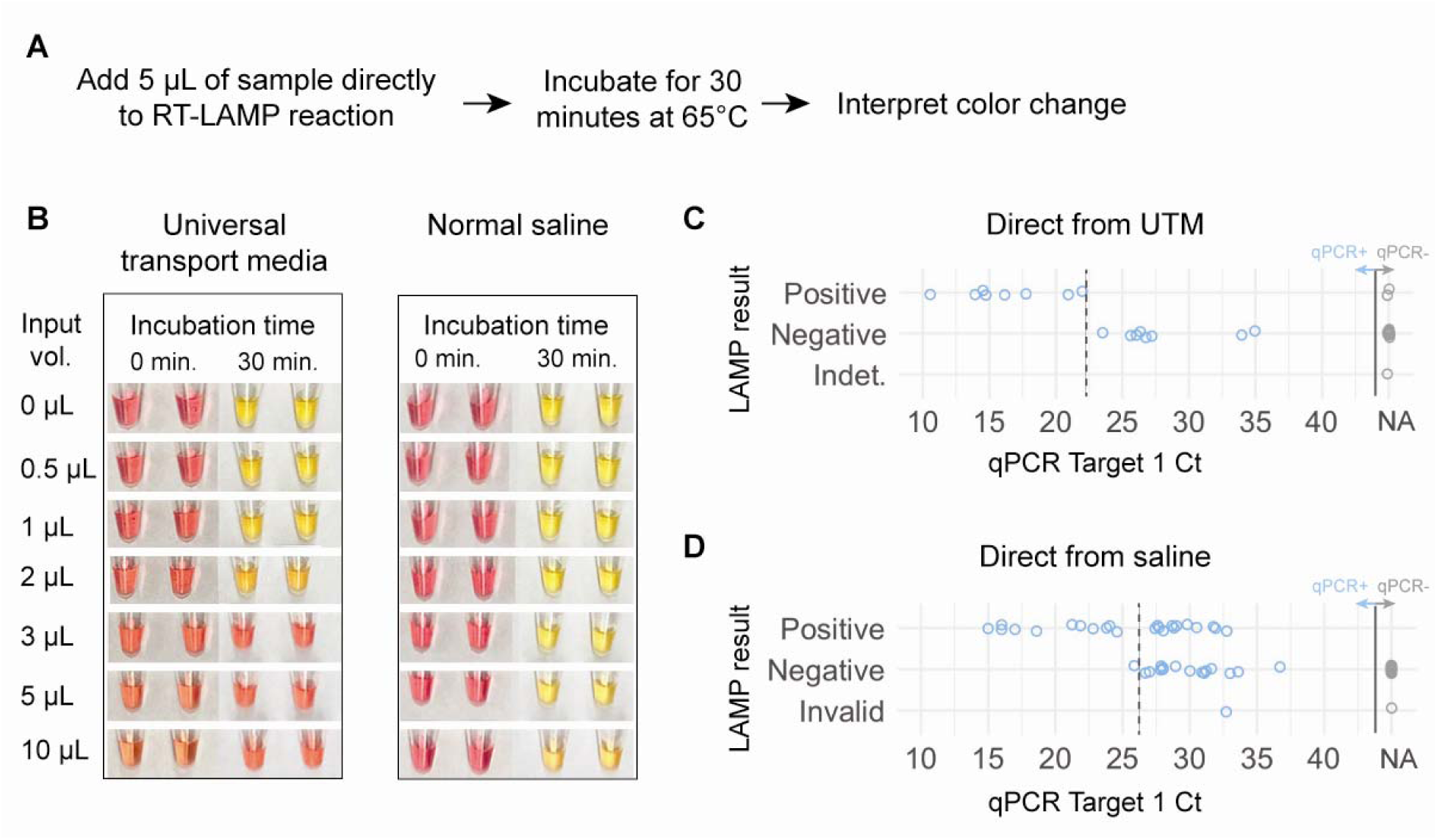
Detection of SARS-CoV-2 directly from nasopharyngeal samples collected in universal transport media (UTM) or 0.9% normal saline. A) Schematic for the thirty-five minute protocol of direct-from-sample testing. B) Determination of the optimal sample input volume for UTM and saline using a standardized 1,000 copy/μL synthetic SARS-CoV-2 input. Samples are pictured before and after the thirty-minute amplification step. C) Comparison of the sensitivity of qPCR to RT-LAMP with direct addition of 1 μL of clinical NP specimen collected in universal transport media (16 qPCR-positive samples, 17 qPCR-negative samples) and testing using the N gene primers alone. D) Comparison of the sensitivity of qPCR to RT-LAMP with direct addition of 5 μL of clinical NP samples collected in saline (40 qPCR-positive samples, 45 qPCR-negative samples), using both the N gene and ORF1a primer sets. One invalid result occurred from a sample that had a negative human actin control and was noted to be bloody. The approximate clinical limit of detection is shown with a dotted line. NA denotes the lack of amplification.

### Direct-from-sample detection

We next asked whether SARS-CoV-2 could be consistently detected from unprocessed clinical UTM and saline samples. When compared to qPCR on an FDA EUA approved platform, the sensitivity of RT-LAMP performed with the SARS-CoV-2 N gene and human actin gene primer sets and direct addition of a UTM specimen was only 50% (Fig. 2C). RT-LAMP could only detect UTM samples with a cycle threshold less than 23, corresponding to approximately 3,000 copies/μL in internal validation studies. There were two false positives, possibly related to interpretation difficulties due to a limited dynamic color range and higher background of the N gene primer set.

We next tested NP specimens directly inoculated into saline transport media using both the N and ORF1a primer sets and the interpretation criteria listed (Table 1). When saline was used as the transport media, we consistently detected samples with cycle thresholds less than 25 and as high as 32, yet the assay sensitivity was only 59% (Figure 2D). Ten samples were detected with both primer sets, four with only the N gene primers, and two with only the ORF1a primers. Among 45 COVID-negative saline samples tested, the color changes were crisper and easier to interpret compared to UTM and there were no false positives. This was consistent with *in silico* and *in vitro* analyses that did not demonstrate cross reactivity between the ORF1a and N gene primer sets and other coronaviruses or respiratory viruses (Supplemental Table 1). Overall, normal saline appeared to be a more amenable sample collection media compared to UTM but direct sample addition to the RT-LAMP reaction remained too insensitive for routine clinical use.

### Assay performance with sample inactivation

We next tested whether the assay sensitivity would improve with a simple inactivation step consisting of TCEP/ EDTA addition to neutralize endogenous RNase activity and heat to release the viral RNA contained within virions and human cells (Fig. 3A). The inactivation step appeared highly effective in nasopharyngeal specimens spiked with serially diluted SARS-CoV-2 control RNA (Supplemental Fig. 1) and enabled performance of limit of detection (LoD) studies. As little as 25 copies/μL of control SARS-CoV-2 RNA could be detected in all 20 replicates using the ORF1a primers, and the N gene primers appeared slightly less sensitive (Fig. 3B).

**Figure 3.**
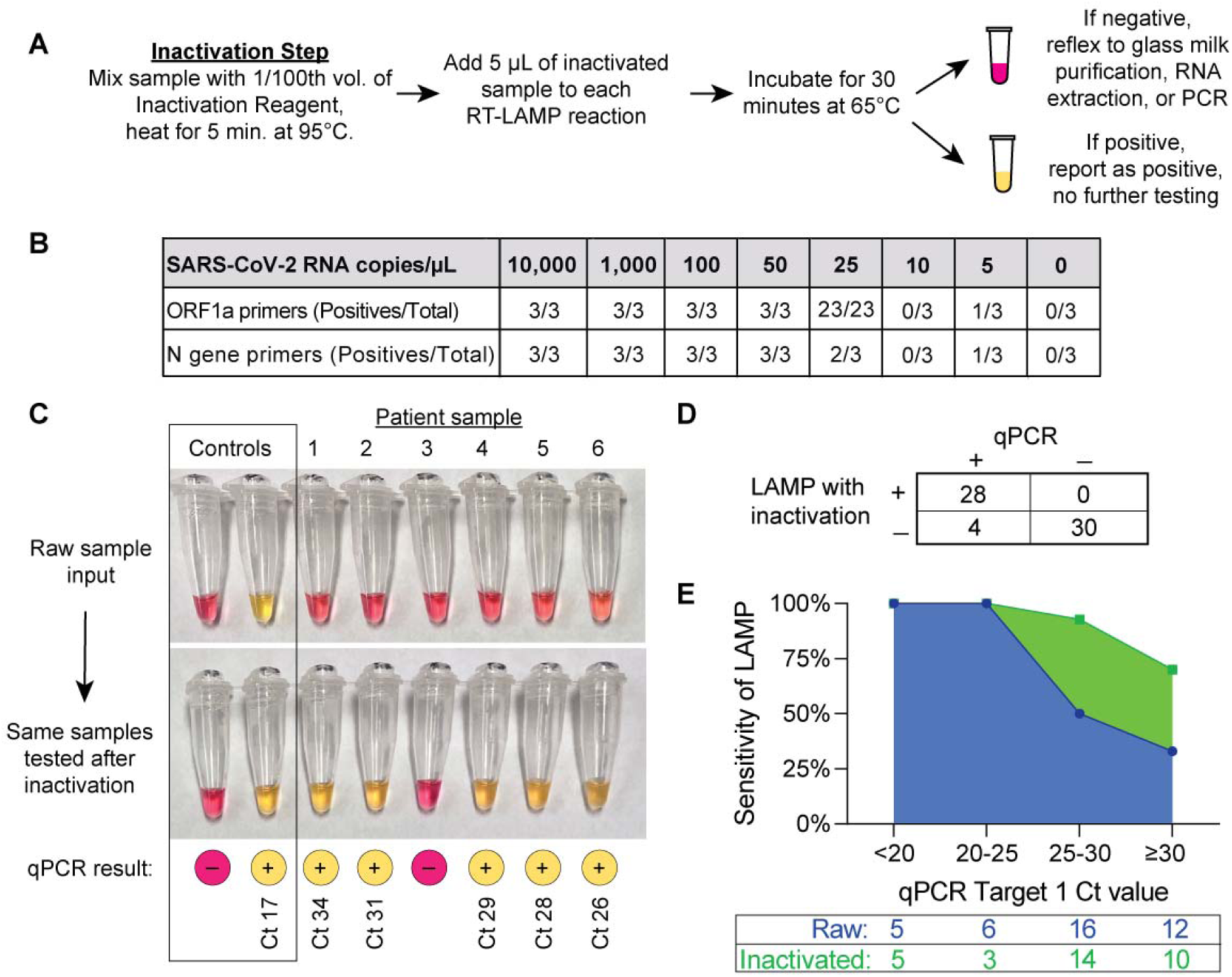
Detection of SARS-CoV-2 after inactivation of RNases and heat lysis from clinical nasopharyngeal swabs collected in saline. A) Schematic for a forty-minute rule-in protocol where samples are first treated with a reducing agent tris(2-carboxyethyl)phosphine (TCEP) and heated to 95 °C for five minutes prior to sample addition to the RT-LAMP reaction. B) Determination of the analytic sensitivity of the RT-LAMP assay with each primer, as determined by synthetic SARS-CoV-2 RNA spiked into inactivated COVID-negative nasopharyngeal samples collected in saline. C) Representative clinical samples illustrating the improvement of RT-LAMP sensitivity after inactivation, with corresponding qPCR results and cycle threshold (Ct) values. Amplification reactions using the ORF1a primer set are shown. D) Overall performance of RT-LAMP in 62 inactivated clinical samples. RT-LAMP results were categorized as positive or negative using the criteria outlined in Table 1. E) Sensitivity of RT-LAMP with or without inactivation as a function of the input SARS-CoV-2 RNA concentration, as determined by qPCR. The total number of samples tested within each Ct range is shown in the table below.

We then incorporated the inactivation step into the testing of clinical samples and observed a substantial improvement in the assay sensitivity. COVID-positive samples that were previously falsely negative with unprocessed sample addition were subsequently positive with both SARS-CoV-2 primer sets after inactivation (Fig. 3C). Importantly, COVID-negative samples remained negative after inactivation (Fig. 3C, Sample 3). To systematically test the efficacy of inactivation, we repeated the assay using inactivation with the available 32 COVID-positive and 30 COVID-negative samples that had originally been tested by direct sample addition. We found 100% specificity and 87.5% sensitivity with this sample set (Fig. 3D). In addition, we found that inactivation enabled the detection of over 95% of samples with a cycle threshold below 30, corresponding to about 40 viral copies/μL, and could detect SARS-CoV-2 in samples with cycle thresholds as high as 33.5 (Fig. 3E). Thus, the combination of a very simple inactivation step followed by RT-LAMP provided a robust rule-in test for SARS-CoV-2 with a sample to result time of approximately forty minutes and minimal labor.

### Assay performance after sample purification

Finally, we asked whether increasing the effective sample input volume using a concentration and purification step could enable detection of samples with very low levels of virus that would otherwise be falsely negative with inactivation alone (Fig. 4A). We used the glass milk protocol developed by Rabe and Cepko [8] to concentrate up to 500 μL of sample into a single RT-LAMP reaction. Purification improved the LoD by ten-fold (Fig. 4B). Two of the four clinical samples that were falsely negative by inactivation alone were positive after the addition of a purification step. The two remaining specimens that tested negative after the purification step may have been truly qPCR-negative due to undergoing several freeze-thaw cycles, but insufficient material remained for repeat testing. All twenty qPCR-negative samples tested negative after purification (100% specificity; Fig. 4C). Overall, purification improved the assay sensitivity by increasing the effective sample input volume.

**Figure 4.**
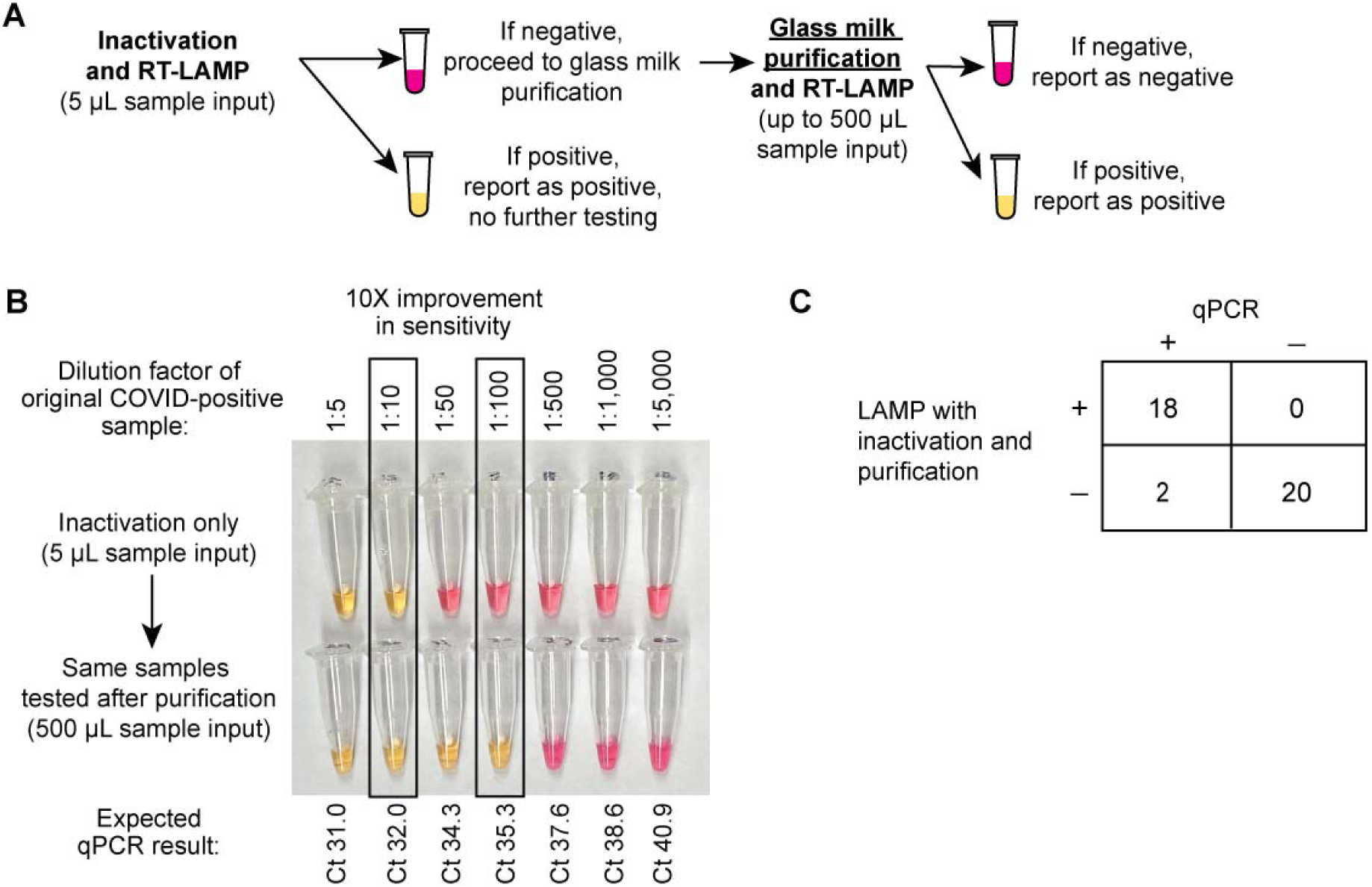
Further improvement in sensitivity by reflexing RT-LAMP-negative inactivated samples to a glass milk purification procedure. A) Schematic for combining inactivation with glass milk purification. B) Demonstration that purification improves the assay detection limit by approximately ten-fold, using a serially diluted COVID-positive patient’s nasopharyngeal specimen. Using the ORF1a primer set, the lowest dilution detected from the inactivated sample was 1:10. After purifying 500 μL of each dilution, the 1:100 dilution could be detected by the same primer set. C) Overall performance of RT-LAMP in 40 clinical samples that were inactivated and then purified, including all four specimens that were falsely negative with inactivation alone.

## Discussion

Here we have demonstrated a simple and inexpensive loop-mediated isothermal amplification assay for the detection SARS-CoV-2 that achieves 87.5% overall sensitivity and 100% specificity with the inclusion of an upfront, five-minute sample inactivation step. Performing an additional glass milk purification step resulted in increased sensitivity of the assay. This assay can be performed in any clinical laboratory or even ad hoc settings, like a mobile laboratory, as it does not require any specialized equipment or highly trained laboratory personnel. Since the required reagents are easily manufactured by multiple manufacturers, access to this test does not rely on traditional commercial diagnostic supply chains that have hindered the broad distribution of SARS-CoV-2 testing. In addition, there is no need for de novo manufacturing. The reagents needed for this assay can be purchased by any laboratory from a number of sources, except for the colorimetric RT-LAMP master mix where the manufacturer has large-scale production in place with millions of reactions worth of product available. We estimate an overall per-sample cost approximately 6 USD. Personnel and overhead costs will also contribute and will vary greatly depending upon the setting. There are essentially no fixed equipment costs. We believe this forty-minute sample-to-answer assay addresses a pressing need for COVID-19 diagnostics worldwide.

Sample preparation is often a time consuming and expensive step of the testing process. We have demonstrated that RT-LAMP can be performed directly from a nasopharyngeal sample but that the assay sensitivity increases by 30% with chemical RNase inactivation using TCEP/EDTA and heat-mediated lysis. In addition to improving assay performance by 30%, the inactivation step described here likely reduces the infectivity of the sample as well [10], reducing the risk of exposure for laboratory personnel. The assay’s sensitivity is further improved by glass milk purification, which is both extremely inexpensive (7 cents per sample) compared to commercial RNA extraction kits (approximately 5 USD per sample) and can be performed without a microcentrifuge, enabling its use in low-resource laboratory environments.

We foresee this assay being using in two ways. The first is a 40-minute rule-in test that uses inactivation followed by RT-LAMP. If the sample is positive and controls are valid, the test is reported as positive for SARS-CoV-2. If it is negative, the sample can then be reflexed to a qPCR-based test or to a glass milk purification, by adding the inactivated sample directly to the binding solution and glass milk. In resource-limited settings that do not have access to on-site qPCR-based diagnostics and the personnel to perform glass milk purification, positive results with the rule-in test would provide a fast turn-around-time and enable effective infection control practices and clinical management, while negative tests would be sent to a reference lab for confirmation by more sensitive detection methods.

While this clinical validation is focused on nasopharyngeal swabs, which are recommended by the U.S. Centers for Disease Control as the most sensitive specimen type for SARS-CoV-2 detection [11], the same methods can be applied to other sample types as well, possibly including saliva [8]. Oropharyngeal specimens are likely to be compatible given their similar composition to nasopharyngeal specimens. Sputum is generally a challenging sample type due to high viscosity and heterogeneity, but we expect the TCEP/EDTA chemical inactivation step to mimic the current recommendation to pre-treat sputum with dithiothreitol, an alternative reducing agent to TCEP [12]. Further clinical studies to assess the range of compatible sample types are underway.

There are several limitations of this assay. The assay is qualitative and does not provide a semi-quantitative cycle threshold number. Additionally, the visual interpretation affords substantial flexibility, but it can also be prone to user errors. Objective color measurements can be performed by measuring the absorbance at 432 and 560 nm, or potentially with a smartphone application. It is critical to use the positive and negative controls as interpretative aids to avoid misinterpreting an orange intermediate color change as positive. The N gene primer set is more likely to give subtle background color changes and additional primer sets will be tested in the future. Like any nucleic acid amplification test, systems must be in place to avoid environmental and sample contamination with post-amplification products. One such precaution is to refrain from opening the reaction vessel after amplification; reactions should be discarded or transferred to a sealed container for later reference, since the color change remains stable for days to weeks. While the assay generally requires very little infrastructure, the operator must abide by laboratory biosafety guidelines and the procedure is most safely performed within a biosafety class II cabinet, although we recognize in many setting this may not be possible [13]. Additionally, the RT-LAMP master mix currently requires storage at -20 °C, which is not ideal for low-resource or remote settings, but this may be ameliorated by lyophilization.

In summary, we present the implementation of a simple RT-LAMP assay for the detection of SARS-CoV-2 that achieves a high sensitivity and specificity in a challenging clinical sample set obtained during the peak of the Spring 2020 COVID-19 pandemic. Future work includes further improving the assay sensitivity through modifications in the primer design, and reaction conditions, creating synthetic positive and negative controls, validating additional sample types, eliminating the cold chain requirement through reagent lyophilization, and applying for FDA emergency use authorization to enable this assay to be used a stand-alone test in a variety of settings.

## Materials and Methods

### Clinical sample collection, qRT-PCR, and storage

Nasopharyngeal samples were collected in 1 mL of sterile physiologic saline from the inpatient units and the Emergency Department (ED) of Massachusetts General Hospital (MGH) between March and April 2020. This study was approved by the Partners Human Research Committee at the Massachusetts General Hospital. The inpatient samples were a convenience set obtained from patients whose COVID-19 status was known (20 qPCR positive, 17 qPCR negative). The ED samples were collected from patients who presented within a 24-hour period and required clinical COVID-19 testing (22 qPCR positive, 45 qPCR negative). In addition, the nasopharyngeal samples collected in 3 mL of universal viral transport media were obtained from excess material collected for routine clinical care.

Upon receipt in the laboratory, samples were tested with an FDA EUA-approved quantitative real-time PCR method (CDC assay-based lab-developed test, Roche SARS-CoV-2 test for the cobas® 6800 system, or, rarely, Cepheid Xpert® Xpress SARS-CoV-2 test). The cobas® 6800 system’s cycle threshold tends to be within 2 cycles of the LDT’s. If the Ct of a saline specimen was not available, the Ct from the paired UTM specimen that was collected simultaneously was used as a proxy. Though these qPCR assays are not truly quantitative due to variability in sample input, approximate conversions between cycle thresholds and viral copies/μL were calculated with a standard curve generated on the LDT by spiking 0, 10^1^, 10^2^, 10^3^, and 10^4^ copies/μL of SARS-CoV-2 N gene RNA into COVID-negative nasopharyngeal specimens. Samples were aliquoted and quickly frozen at -20 °C for additional testing to avoid RNA degradation. There was no noticeable difference in RT-LAMP assay performance between fresh and frozen samples.

### Control SARS-CoV-2 RNA

The synthetic SARS-CoV-2 RNA control used for spike-in experiments was obtained from Twist Bioscience (MT007544.1, 1×10^6^ RNA copies/μL) and was freshly diluted as needed into nuclease-free water.

### RT-LAMP primers

The SARS-CoV-2 ORF1a gene [8], SARS-CoV-2 N gene [9], and human actin B gene (generously provided by New England Biolabs) primer sequences are listed in Supplemental Table 2. The ORF1a primers were combined into a 10X primer stock using 16 μM of Forward Inner Primer (FIP), 16 of Backward Inner Primer (BIP), 2 μM of F3, 2 of B3, 4 of Forward Loop (LF), and 4 μM of Backward Loop (LB). The N gene and human actin primer stocks were consisting of the same primer proportions at a 25X concentration.

### RT-LAMP assay

RT-LAMP testing was performed in biosafety level 2, CLIA-certified laboratory space. Each 25μL RT-LAMP reaction was performed as described by the manufacturer’s protocols with WarmStart® Colorimetric RT-LAMP 2X Master Mix (New England Biolabs, M1800) using a 1 μL sample input for samples collected in UTM and 5 μL input for samples collected in normal saline. The interpretative criteria are listed in Table 1. After interpretation, tubes were discarded or stored in sealed bags without re-opening to prevent post-amplification contamination of workspaces.

### Inactivation and purification procedures

The 100X inactivation reagent and purification reagents were prepared as described elsewhere [8]. The inactivation reagent was comprised of 0.25 M of Tris(2-carboxyethyl)phosphine hydrochloride (TCEP-HCl; Millipore Sigma, 580567), 0.1 M RNase-free EDTA (ThermoFisher Scientific, AM9260G), and 1.1 N NaOH, diluted in UltraPure water (ThermoFisher Scientific, 10977015). The saline NP sample was mixed with 1/100^th^ volume of the 100X TCEP/EDTA-based inactivation reagent and heated at 95 °C for 5 minutes. The sample was then cooled on ice and directly added to the RT-LAMP reaction or used for purification. When purification was performed, 250-500 μL of the inactivated sample was mixed with 5 μL of glass milk in a 1.5 mL tube, thoroughly resuspended, and mixed with half the initial sample volume of binding reagent. As described elsewhere [8], the binding reagent was comprised of NaI (Millipore Sigma, 793558), HCl (Millipore Sigma, 320331), and Triton X-100. The sample was then incubated at room temperature for 10 minutes with manual inversions approximately every two minutes to resuspend the silica. The samples were briefly spun in a mini benchtop centrifuge for several seconds and the supernatant was poured off. The pellet was washed with 700 μL of 80% ethanol and briefly spun. The supernatant was poured off again and briefly respun. Any visibly remaining ethanol was removed with a P20 pipette and the pellet was air-dried on a heat block at 65 °C for five minutes or until the pellet was visibly dry. Twenty-five μL of colorimetric RT-LAMP reaction mix was added to the pellet, resuspended, and transferred to a 0.2 mL tube for incubation at 65 °C for 30 minutes and visual inspection.

### Limit of detection

An initial LoD study was performed for each SARS-CoV-2 primer by spiking-in serially diluted synthetic SARS-CoV-2 RNA into an inactivated, COVID-negative nasopharyngeal saline media. Five microliters of sample were tested in triplicate at final concentrations of 10^4^, 10^3^, 100, 50, 25, 10, 5, and 0 copies per μL. An additional twenty replicates were performed at the concentration predicted to be the LoD, as defined by the FDA at the lowest concentration at which 19/20 replicates are positive. The purification dilution experiments were performed by making serial dilutions of a COVID-positive sample in COVID-negative nasopharyngeal specimens.

### Cross-reactivity

Cross-reactivity of the N gene primer set with SARS-CoV-1 and MERS were assessed with plasmid controls (Integrated DNA Technologies, 10006624 and 10006623). Plasmids containing the ORF1a region of SARS-CoV-1 and MERS were not available, thus *in vitro* testing of ORF1a primers with SARS-CoV-1 and MERS were not be performed. In addition, to assess for primer cross-reactivity with common respiratory pathogens, the assay was performed on ten clinical samples collected prior to the outbreak of SARS-CoV-2 (February through April 2019) that were known to contain a respiratory virus due to clinical multiplexed PCR testing (Biofire Film Array Respiratory Panel, RP2). Since the samples were collected in UTM, RNA was first extracted from 140 μL of nasopharyngeal swab UTM using a Qiagen Viral RNA Mini Kit (Qiagen, 52906) and eluted in 60 μL, and 5 μL of extracted RNA was added to each RT-LAMP reaction.

## Data Availability

Data available within the article or its supplementary materials.

## Acknowledgements

New England Biolabs generously provided the N gene and human actin primer sets and initial master mix. Maria Vareschi, John Texeira, and MGH Emergency Department (ED) staff enabled the ED sample collection. Dr. Tyler Miller and MGH nursing staff enabled the inpatient saline sample collection. Dr. Hetal Marble provided insight into the development of the assay into a commercially available kit. Dr. Virginia Pierce and Dr. Sarah Turbett provided feedback and invaluable guidance into the clinical validation process. Last but certainly not least, the incredibly dedicated MGH microbiology technologists performed the qPCR assays as part of their daily excellence in clinical care.

